# Performance and safety of induced sputum procedure in young children in Malawi: a prospective study

**DOI:** 10.1101/2021.08.12.21261823

**Authors:** Wongani WN Nyangulu, Herbert HT Thole, Angella AC Chikhoza, Mike MM Msakwiza, James JN Nyirenda, Mphatso MC Chisala, Pui-Ying P-YIT Iroh Tam

## Abstract

**Background:** Collecting sputum specimens are a challenge in infants and young children. We assessed performance and safety of induced sputum (IS) collection in this population, embedded in a prospective study evaluating respiratory cryptosporidiosis in Malawian children with diarrheal disease.

**Methods:** We assessed sputum quality and correlation with detection of cryptosporidium, and evaluated safety and adverse events in 162 children.

**Results:** Among 159 stool specimens tested, 34 (21%, 95% CI 15.0 – 28%) were positive for *Cryptosporidium* spp. There were160 IS and 161 nasopharyngeal (NP) specimens collected. The majority of IS specimens 122/147 (83%) were clear in appearance, and 132/147 (90%) were of good quality. Among the respiratory specimens tested, 10 (6.3%, 95% CI 2.5 - 10) IS and 4 (3% (95% CI 0 – 5)) NP were positive for *Cryptosporidium* spp. When stool cryptosporidium PCR was the gold standard, IS PCR sensitivity was higher (29 %, 95% CI 22 – 37) compared to NP PCR (12%, 95% CI 7 – 17) for detection of *Cryptosporidium* spp. One (0.4%) adverse event occurred, a drop in oxygen saturations at 30-minute post procedure evaluation. Consciousness – level, median respiratory rate and oxygen saturations were unchanged, before or after IS.

**Conclusions:** IS provides good quality specimens, is more sensitive than NP specimens for diagnosis of respiratory cryptosporidiosis, and collection can be done safely in children hospitalized with diarrheal disease.

## Introduction

Sputum specimens are routinely collected for diagnosis of respiratory diseases in adults (1). However, this is more challenging in infants and young children (1) who require sputum induction and/or gastric aspiration to obtain diagnostic specimens (2). Children have difficulty producing an adequate specimen, difficulty with expectoration, a tendency to swallow sputum and have high nasopharyngeal (NP) carriage of potential contaminants, including *Streptococcus pneumoniae* (19 – 38%), *Haemophilus influen*zae (13 – 25%), *Moraxella catarrhalis* (22 – 39%) and *Staphylococcus aureus* (16 – 36%) (1, 3, 4).

In Malawi, gastric aspiration is the standard method for obtaining pediatric sputum specimens in public hospitals. These facilities are usually under resourced, lack equipment for induced sputum (IS) collection and staff trained to conduct IS. IS procedures are mostly done in research settings. However, IS has better diagnostic yield for pulmonary tuberculosis compared to gastric aspiration (5, 6). It is also safe and well tolerated with minimal adverse events after the procedure, even in severely ill children (7, 8). Few studies evaluating safety of IS were done in low resource settings (8, 9), and none have been done in a public tertiary hospital in Malawi.

In this study, we assess performance and safety of IS collection in infants and young children enrolled in a prospective study evaluating for respiratory cryptosporidiosis in children hospitalized with diarrheal disease (10).

## Materials and methods

### Participants

Participants were children 2 – 24 months of age hospitalized with primary diagnosis of diarrhea as part of a study evaluating the role of respiratory cryptosporidiosis in pediatric diarrheal disease (Iroh Tam et al, manuscript under review). Details of this study are published elsewhere (10). All enrolled patients had IS, NP and stool specimens collected for *Cryptosporidium* spp. testing at enrollment, and then every two weeks up to eight weeks post-enrollment if any of the enrollment specimens were positive.

### Specimen collection

IS was only done in patients without any contraindications as described in the published protocol. IS was done as follows: patients were given nebulized salbutamol followed by 3% sodium chloride mist inhaled for 5-15 minutes. A sterile catheter connected to a suction machine was passed through the nose into the posterior nasopharynx. Suction was applied only once to aspirate contents and immediately stopped. The catheter was withdrawn and flushed using sterile normal saline. IS was stopped if the child could not tolerate the procedure or if there was an adverse reaction. The child was followed up at 30 minutes, 2 hours and 4 hours post-procedure to monitor for any adverse events.

### Laboratory procedures

Sputum quality was assessed at enrollment by microscopy, noting the appearance (clear v. cloudy) and counting the number of squamous epithelial cells (SECs) per low power field (LPF). Good quality sputum was classified as ≤ 10 SECs per LPF. The number of neutrophils per LPF were also recorded. PCR testing was done to detect *Cryptosporidium* spp., using methods described elsewhere (10).

### Statistical analysis

Data was analyzed using STATA 13 (Stata Corp, USA). We reported proportions for all categorical variables, and median and interquartile range for non-normally distributed continuous variables. Performance was measured by calculating sensitivity and specificity of PCR using stool and sputum cryptosporidium PCR, respectively, as the gold standards. Results are reported at the 5% significance level.

## Results

A total of 162 children were enrolled into the study. Of these, 28 had more than one IS done, and a total of 263 IS procedures were carried out. The median age of enrolled children was 11 months (interquartile range 8-14 months), and 95 (59%) were male. Table 1 shows the health status of children before IS procedure.

**Table 1.** Health status of children before and after induced sputum procedure

| Characteristic | Immediately before | Immediately after |
| --- | --- | --- |
| Fever (Temperature > 37.5 °C; %) | 18 (11) | ND |
| Oxygen saturations (%), median (IQR) | 99 (98 – 100) | 98 (98 – 99) |
| Respiratory rate (bpm), median (IQR) | 35 (32 – 38) | 36 (34 – 38) |
| Looking sick requiring referral to the hospital (%) | 52 (32) | ND |
| Looking unwell uncomfortable (%) | 52 (32) | ND |
| Looking comfortable well and playful (%) | 58 (36) | ND |
| Receiving supplemental oxygen via nasal prongs (%) | 3 (2) | 3 (2) |
| Receiving intravenous fluid (%) | 38 (24) | ND |
| Bipedal edema | 5 (3) | ND |
| Consciousness level (AVPU* score) |  |  |
| 1. No. (%) V, P or U | 0 (0) | 0 (0) |
| 2. No. (%) with any decrease from baseline | ND | 0 (0) |

\*A = alert, V = verbal, P = pain, U = unresponsive; ND = Not Done

There were 147 induced sputum specimens with documented appearance and cell counts. The majority of sputum specimens 122/147 (83%) were clear, and only a few were either cloudy or mucoid (16% and 1% respectively). Most of the specimens 132/147 (90%) were of good quality with < 10 SECs per LPF. Only one specimen had > 10 neutrophils per LPF. None of these characteristics was significantly associated with *Cryptosporidium* spp positivity (Supplementary material).

Among 159 stool specimens tested at enrollment, 34 (21% (95% CI 15 – 28%)) were positive for *Cryptosporidium* spp. Among 160 IS and 161 NP specimens that were tested 10 (6% (95% CI 3 – 10)) and 4 (3% (95% CI 0 – 5)) respectively were positive. When stool cryptosporidium PCR was used as the gold standard, sensitivity of IS PCR compared to NP PCR for detection of *Cryptosporidium* spp was 29 % (95% CI 22 – 37) compared to 12% (95% CI 7 – 17). Both tests had specificity of 100% (95% CI 100 – 100). When sputum PCR was used as the gold standard, NP PCR had sensitivity of 40% (95% CI 32 – 48).

There was one adverse event following all IS procedures representing a risk of 0.4%. A child experienced drop in oxygen saturations below 92% at 30-minute evaluation post IS. The child did not present with any respiratory symptoms at hospitalization. The patient was managed supportively and oxygen saturations at subsequent follow up intervals were normal without any intervention.

Median oxygen saturations and respiratory rates remained stable before and after IS. There was no change in the number of children requiring supplemental oxygen immediately before IS to 4 hours after the procedure. There were 9 children (6%) who had tachypnea immediately before IS. Immediately after IS, 8 children (5%) had tachypnea. At 30 minutes, 2 hours and 4 hours post procedure, 5 (3%), 3 (2%) and 6 (4%) children had tachypnea. All episodes resolved spontaneously. No children experienced decreased level of consciousness before or after IS.

There were 3 children who were on oxygen therapy before the procedure. However, their oxygen saturations were above 92% before and after IS procedure.

## Discussion

This is one of the few studies comparing performance of IS to NP for the detection of respiratory parasites, and one of the first done in a low resource setting.

Other studies of IS have established the procedure to provide good quality sputum specimens with high microbial yield in children (4). In a prospective study in South Africa, IS was done in 142 children. Compared to gastric lavage, yield for *Mycobacterium tuberculosis* was 4% higher for IS specimens (95% CI 0 – 5.6, p-value 0.08) (11). In another study in The Netherlands of children admitted with acute lower respiratory infection, 98 children had IS done. A total 89/98 (91%) had good quality sputum, and bacterial pathogens were isolated from 22/89 (25%) (12). In India, IS was done in 120 children diagnosed with pneumonia. There was good quality sputum in 64 children (53.3%). Among these, 45/64 (70%) had isolates of *Klebsiella pneumoniae* (38.2%) and *Streptococcus pneumoniae* (14.8%) and other bacteria (9).

IS procedure in young children hospitalized with diarrheal disease was safe and well tolerated in our study. Adverse events were rare and involved only one subject (0.4%), and this is consistent with studies done elsewhere including the PERCH study, where the proportion of adverse events was also low at 0.34% (8), even though that study recruited children with moderate to severe pneumonia. The PERCH study team recommended safety monitoring during and after IS up to two hours post procedure with close attention to oxygen saturations in severely ill children with pneumonia (8). PERCH had a larger sample size and recorded more adverse events than our study. Therefore, our findings do not support the need for post procedure follow up, though it is important to note that we did not recruit and conduct IS on children with a primary respiratory diagnosis.

We demonstrated that well trained staff in public hospitals can conduct IS with good sputum quality for diagnosis of respiratory pathogens. We also demonstrated that this procedure is safe in a low resource setting. This has implication on practice in the wards where IS is not regularly done due to lack of trained staff and materials including hypertonic saline. Medical and nursing staff can be trained to conduct IS safely with good diagnostic yield and materials required for the procedure are relatively inexpensive. The low rate of adverse events among patients with a primary diarrheal diagnosis indicates that clinical protocols that emphasize close monitoring of patients after the procedure is unnecessary.

The procedure was also acceptable to parents and guardians of children admitted in hospital. In our study, none of the parents were familiar with the IS procedure. All were worried about the procedure, thought it was uncomfortable and would cause pain. However, after detailed explanation of the procedure, all parents provided consent. Those who had repeat procedures done did not express similar misgivings towards IS during the follow up study visits.

The main limitation of our study is a smaller than anticipated sample size. Based on the expected sample size for the main study and expected cryptosporidium positivity rate, we expected to conduct over 300 IS procedures (10). The emergence of the Severe Acute Respiratory Syndrome-Coronavirus 2 (SARS-CoV-2) global pandemic resulted in suspension of all study activities.

## Conclusions

In summary, we demonstrated that IS procedure in young children hospitalized with diarrheal disease provides higher sensitivity compared to NP swabs for diagnosis of respiratory cryptosporidiosis, and is safe and acceptable in a low resource setting. Our study and these findings suggest that IS can perform well and be done safely in our setting for diagnosis of respiratory pathogens.

## Data Availability

Data will be made available upon reasonable request to the primary study principal investigator (P-YIT).

## Authors’ contributions

WN and P-YIT conceived the study. WN, HT, AC and MC collected data and specimens. WN performed statistical analyses. WN and P-YIT drafted the original manuscript. All authors reviewed and approved the final manuscript.

## Acknowledgements

We would like to thank the staff and patients at the department of paediatrics for allowing us to conduct this study in the paediatrics wards; David Moore and Tanja Adams with training the CryptoResp clinical and laboratory teams; and staff at Malawi Liverpool Wellcome Trust Clinical Research Programme for providing logistical and material support during the study period.

## Ethical approval

The College of Medicine Research Ethics Committee (COMREC) (P.07/18/2438) and the Liverpool School of Tropical Medicine Research Ethics Committee (18-066) approved the main study.

