## Supplementary materials for "Performance and safety of induced sputum procedure in young children in Malawi: a prospective study"

Section one: Analysis of association between sputum characteristics and cryptosporidium positivity

Table one – A 2 x 2-table showing cryptosporidium positivity by sputum appearance

|  | Appearance | | |  |
| --- | --- | --- | --- | --- |
| Cryptosporidium positivity | Clear | Cloudy | Mucoid | Total |
| Negative | 115 | 22 | 2 | 139 |
| Positive | 7 | 1 | 0 | 8 |
| Total | 122 | 23 | 2 | 147 |

Table two – A 2 x 2-table showing cryptosporidium positivity by sputum neutrophil count

|  | Neutrophils | | |  |
| --- | --- | --- | --- | --- |
| Cryptosporidium positivity | < 10 | 10 – 25 | - 25 | Total |
| Negative | 135 | 3 | 1 | 139 |
| Positive | 8 | 0 | 0 | 8 |
| Total | 143 | 3 | 1 | 147 |

Table three – A 2 x 2-table showing cryptosporidium positivity by sputum epithelial cell count

|  | Epithelial cells | | |  |
| --- | --- | --- | --- | --- |
| Cryptosporidium positivity | < 10 | 10 – 25 | - 25 | Total |
| Negative | 125 | 9 | 5 | 139 |
| Positive | 8 | 0 | 0 | 8 |
| Total | 133 | 9 | 5 | 147 |

Table four – A summary of Pearson chi – squared test of association between cryptosporidium positivity and sputum characteristics

| Characteristic | Pearson chi – squared statistic | P - value |
| --- | --- | --- |
| Appearance | 0.1893 | 0.910 |
| Neutrophils | 0.2367 | 0.888 |
| Epithelial cells | 0.8906 | 0.641 |

Table five – A summary of univariate logistic regression analysis with cryptosporidium positivity (0 = negative, 1 = positive) as the outcome variable

| Characteristic | Unadjusted OR | P - value | 95% Confidence Interval |
| --- | --- | --- | --- |
| Appearance   - Cloudy - Mucoid | 0.75  1 (Dropped by STATA) | 0.790  - | 0.87 – 6.37  - |
| Neutrophils   - 10 – 25 - > 25 | 1 (Dropped by STATA)  1 (Dropped by STATA) | -  - | -  - |
| Epithelial cells   - 10 – 25 - > 25 | 1 (Dropped by STATA)  1 (Dropped by STATA) | -  - | -  - |

Section two: Sensitivity and specificity analysis for induced sputum and nasopharyngeal swab procedures

Table one – A 2 x 2-table showing sputum polymerase chain reaction test results using stool specimen as the gold standard

|  | Stool Cryptosporidium PCR as gold standard | |  |
| --- | --- | --- | --- |
| Sputum PCR | Negative | Positive | Total |
| Negative | 124 | 24 | 148 |
| Positive | 0 | 10 | 10 |
|  | 124 | 34 | 158 |

Table two – A summary of sensitivity, specificity, positive predictive value and negative predictive value for sputum PCR

|  | % | 95% Confidence Interval |
| --- | --- | --- |
| Sensitivity | 29.4 | 22.3 – 36.5 |
| Specificity | 100 | 100 – 100 |
| Positive Predictive Value | 100 | 100 – 100 |
| Negative Predictive Value | 83.8 | 78.0 – 89.5 |
| Prevalence | 21.5 | 15.1 – 27.9 |

Table three – A 2 x 2-table showing nasopharyngeal swab polymerase chain reaction test results using stool specimen as the gold standard

|  | Stool Cryptosporidium PCR as gold standard | |  |
| --- | --- | --- | --- |
| NP swab | Negative | Positive | Total |
| Negative | 125 | 30 | 155 |
| Positive | 0 | 4 | 4 |
|  | 125 | 34 | 159 |

Table four – A summary of sensitivity, specificity, positive predictive value and negative predictive value for NP PCR

|  | % | 95% Confidence Interval |
| --- | --- | --- |
| Sensitivity | 11.8 | 6.8 – 16.8 |
| Specificity | 100 | 100 – 100 |
| Positive Predictive Value | 100 | 100 – 100 |
| Negative Predictive Value | 80.7 | 74.5 – 86.8 |
| Prevalence | 21.4 | 15.0 – 27.8 |
